# Algorithm Fairness in Predicting Unmet Preventive Care: Evidence from 16 European Countries using SHARE

**DOI:** 10.1101/2025.09.09.25335389

**Authors:** Toby Kai-Bo Shen, Vincent Cheng Sheng Li, Nick Meng-Huan Chen, Jennifer Sheng Hui Hsu, Rifat Atun, Valerie Tzu Ning Liu, Charlotte Wang, David Bin-Chia Wu, Pin-Chun Yeh, John Tayu Lee

## Abstract

**Background:** Preventive cares are critical to achieve health equity but remain underutilized, particularly among socioeconomically disadvantaged populations. While machine learning (ML) models have shown promise in predicting unmet needs, the fairness and generalizability across national contexts remain poorly understood. This study evaluates the predictive performance and algorithmic fairness of ML models in identifying unmet preventive care needs across 16 European countries.

**Methods:** The study used cross-sectional data of 51,720 adults from Wave 9 of Survey of Health, Ageing and Retirement in Europe (SHARE). We trained and tested ML models including Logistic Regression, Random Forest, XGBoost, LightGBM, Gradient Boosting, DNN and FCN, applied to predict five preventive care outcomes. Model performance was assessed by the area under the receiver operating characteristic curve (AUC). Fairness was evaluated by demographic parity and equalized odds across countries and socioeconomic subgroups. SHAP values quantified the feature importance.

**Results:** LightGBM achieved the highest AUC (0.73–0.81) but exhibited substantial variability across countries (AUC range: 0.53–0.94) and socioeconomic strata. Fairness assessments revealed pronounced disparities, demographic parity differences ranged from 0.0027 to 0.9613 across countries, and inequities were notable among high-income and high education subgroups. Age, income, outpatient visits, and social engagement identified as key predictors.

**Conclusion:** This study provides the evaluations of algorithmic fairness in ML-based prediction of preventive care needs across multiple national contexts. Significant geographic and socioeconomic disparities in model performance highlight the need for localized model calibration and fairness-aware to prevent the reinforcement of health inequities.

## Background

Preventive care services including vaccination, eye examination, colon screening, dental care, and mammograms—are central to achieving Sustainable Development Goal (SDG) 3 by improving health outcomes and reducing health disparities.^1^ ^2^ Despite strong evidence of cost-effectiveness and public health impact, these services remain underutilized across Europe, particularly among older adults and socially disadvantaged populations^3^ ^4^. Identifying individuals at risk of unmet preventive care is crucial for improving health equity and targeting interventions efficiently.^5^

Machine learning (ML) offers significant potential in this context by enabling accurate prediction of unmet health needs based on large-scale, multidimensional data.^6^ Compared to traditional statistical models, ML techniques are more flexible in handling complex interactions, nonlinear relationships, and high-dimensional feature spaces.^7^ Prior studies have demonstrated the utility of ML models in forecasting healthcare utilization and predicting gaps in care, but most have focused on single-country settings without addressing broader generalizability or fairness concerns.^8–10^

A major challenge in applying ML models to population health is ensuring fairness particularly when models are trained on multinational datasets and applied across diverse social and healthcare systems.^11^ ^12^ Europe presents a unique test case due to its substantial variation in health systems, socioeconomic structures, and cultural attitudes toward prevention.^13^ Using a single model across countries without adjustment may introduce systematic biases, reinforce existing health inequities, and misallocate resources.^14–16^

Algorithmic fairness refers to the equitable performance of predictive models across subgroups defined by protected characteristics such as income, education, gender, or region.^17^ In healthcare, fairness is essential to ensure that models do not disproportionately misclassify or overlook vulnerable populations.^18^ ^19^ However, evidence on fairness in ML models for preventive care remains limited particularly in multi-country settings.^20^

To address this gap, we evaluated the performance and fairness of multiple ML models in predicting unmet preventive care needs across 16 European countries using data from the Survey of Health, Ageing and Retirement in Europe (SHARE). We examined both overall predictive accuracy and subgroup fairness using standard algorithmic fairness metrics, focusing on how model performance varies by country and socioeconomic status. Our study provides new insights into the ethical deployment of ML in public health and highlights the importance of localized, fairness-aware model development.

## METHODS

### Dataset

This study analyzed cross-sectional data from Wave 9 (2021) of Survey of Health, Ageing, and Retirement in Europe (SHARE), which provides high-quality information on health, socioeconomic status, and social networks among individuals aged 50 and older. The original dataset encompassed approximately 70,000 interviews from 23 European countries, collected using multistage stratified probability sampling by around 2,000 interviewers. To enhance model performance, we refined the data by removing incomplete responses and excluding 7 European countries with fewer than 200 participants. Our final study sample contains a final sample of 51,720 participants from 16 European countries.

### Outcomes

This study evaluates unmet preventive care needs across five key outcomes, including vaccination, eye examination, mammogram, colon screening, and dental care. Outcomes were assessed based on adherence to guidelines, including flu vaccination for at-risk populations, biennial eye exams, mammograms, colon cancer screenings, and annual dental visits. Using machine learning model to identify patterns of unmet needs also their socio-economic determinants.

### Predictors

A total of 77 features were included across the following domains. A detail description of the features and their definition can be found in Appendix Table:

#### Demographic factors

Age, gender, country, migration, religion, marital status, region, residence type, living alone, household size and number of children.

#### Socioeconomic factors

Education level, economic status, job, paid work, other job, hours of work per week, personal income, household income, home ownership, owe, mortgage.

#### Medical Indicators

Long-term health problem, clinical disease (heart problems, hypertension, cholesterol, stroke, diabetes, lung disease, cancer, ulcer, Parkinson’s, cataracts, fracture, dementia, psycho, arthritis and kidney), near vision, hearing problem, level of pain.

#### Health care utilization

Preventive care, vaccination, eye examination, mammogram, colon screening, dental care, outpatient, inpatient, healthcare facility stays, medicine, polypharmacy, satisfied with health insurance, supplementary health insurance, home care and nursing home.

#### Functional and physical health

Self-report of health, instrumental activities of daily living (IADLs), mental health (EUROD-12), health literacy, limit to work, limit to activity, inactivity, handgrip, hearing aids, cane and wheelchair.

#### Lifestyle and health behaviors factors

Drinking, smoking and consuming of fruits or vegetables.

#### Social and family dynamics

Make ends meet, quality of life (CASP-12), satisfied with life, frequency and number of social activity and voluntary activities.

### Data Processing

In this study, we applied one-hot encoding to preprocess categorical variables with more than two categories. Comprehensive documentation of all preprocessing steps, including the handling of missing data, variable encoding, and feature engineering, is available on GitHub.

Subsequently, the dataset was partitioned into training (80%) and testing (20%) subsets, with the training set utilized for model development and the testing set reserved for performance evaluation. Hyperparameter tuning was conducted using a randomized search approach, incorporating 5-fold cross-validation across 30 iterations to enhance model accuracy and reliability.

### Statistical Analysis

#### Model Development and Fairness Evaluation

This study utilized machine learning models, including Logistic Regression, Random Forest, XGBoost, Gradient Boosting, LightGBM, Deep Neural Networks (DNN), and Fully Convolutional Networks (FCN), with a baseline Logistic Regression model for performance comparison.^21^ Model evaluation metrics such as accuracy, precision, and recall (sensitivity) were employed to assess predictive capability comprehensively. To ensure robustness and generalizability, we applied the K-fold validation method, which involved iterative training and validation on distinct data splits.^22^ Model discrimination was visualized using Receiver

Operating Characteristic (ROC) curves and quantified with Area Under the Curve (AUC) metrics. Additionally, Subgroup Fraction Curves were used to evaluate discrimination across subpopulations. Comparative AUROC plots provided insights into model performance within specific demographic and social determinant subgroups, ensuring a rigorous evaluation of model reliability and fairness across diverse populations. We assessed fairness through demographic parity and equalized odds to ensure that model predictions remain equitable across countries.

### SHAP and Feature Importance Analysis

Feature importance was analyzed across socioeconomic and country subgroups using Shapley Additive Explanations (SHAP), an interpretable machine learning approach to assess the contributions of key feature importance of the machine learning model. SHAP employs a game-theoretic framework to decompose model predictions into additive contributions by aggregating local importance values across all observations, yielding both global rankings and individual-level explanations. We computed SHAP values within each subgroup to assess the consistency of critical predictors, highlighting overlaps and divergences in influential factors across diverse patient profiles and geographic contexts.

### Ethics

The study was approved by the NTU Ethical Review Board (NTU REC-No.: 202411HM030) and adhered to ethical guidelines for the use of secondary data. The use of SHARE Wave 9 data was approved by the Ethics Committee of the Max Planck Society for the Progress of Science. All analysis were conducted using Python (v3.12). All data analysis python code is available for download at: https://github.com/HealthEconomicsandPolicyInnovationLab/SHARE-unmet-preventive-care-needs

## RESULTS

### Study Population

The study included 51,720 adults aged 50 years and older from 23 European countries (excluding 7 European countries with fewer than 200 participants on model developing), distributed in five preventive care services with unmet needs were most prevalent for colon screening (75.01%), followed by vaccination (59.99%), eye examination (52.94%), mammogram (55.92% among eligible women), and dental care (48.56%). Women represented 57.20% of the sample, and 48.88% of participants were classified as low-income subgroup. Educational level was low in 29.85% of participants. The most common chronic conditions were hypertension (28.8%), high cholesterol (16.1%), and diabetes (8.8%). Croatia (9.06%), Estonia (8.67%), and Germany (8.64%) accounted for the highest country representation. Disparities in unmet care were more pronounced among participants with low socioeconomic status and multiple chronic conditions, emphasizing the need for targeted, equity-driven preventive health strategies across diverse European populations.

**Table 1.**
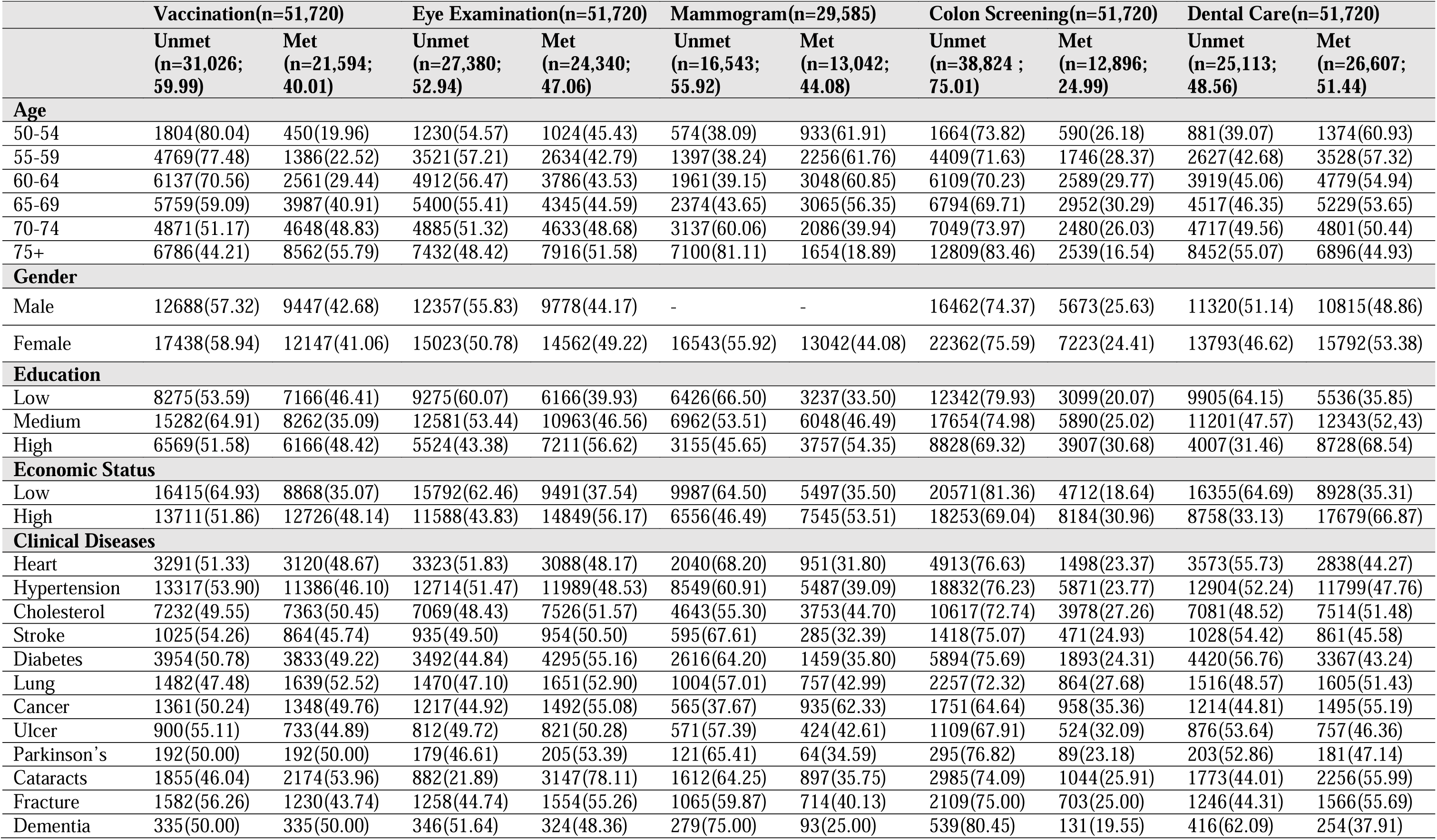
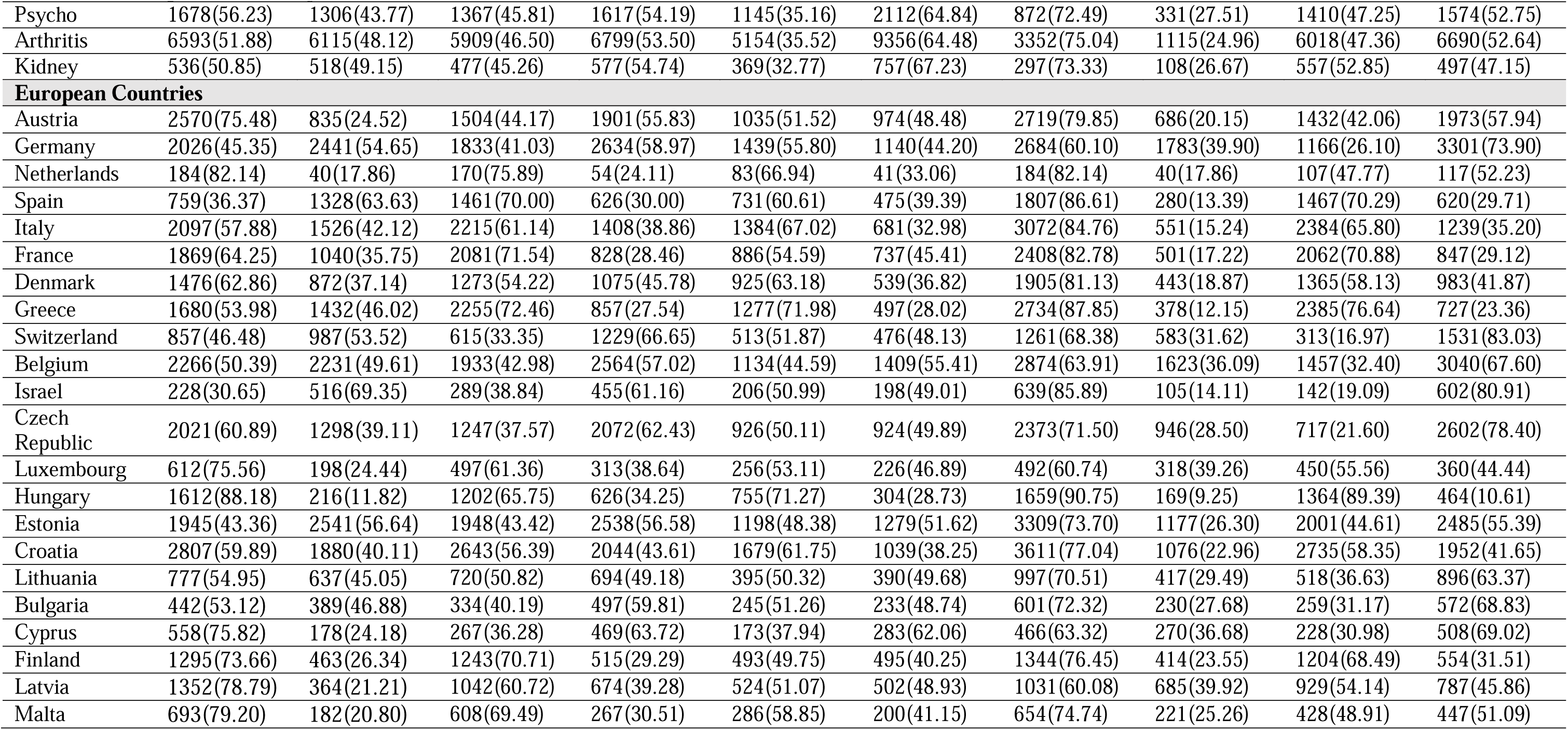
Descriptive summary (n, %)

| **Table 1. Descriptive summary (n, %)** |  |  |  |  |  |  |  |  |  |  |
| --- | --- | --- | --- | --- | --- | --- | --- | --- | --- | --- |
|  | **Vaccination(n=51,720)** |  | **Eye Examination(n=51,720)** |  | **Mammogram(n=29,585)** |  | **Colon Screening(n=51,720)** |  | **Dental Care(n=51,720)** |  |
|  | **Unmet**<br>**(n=31,026;**<br>**59.99)** | **Met**<br>**(n=21,594;**<br>**40.01)** | **Unmet**<br>**(n=27,380;**<br>**52.94)** | **Met**<br>**(n=24,340;**<br>**47.06)** | **Unmet**<br>**(n=16,543;**<br>**55.92)** | **Met**<br>**(n=13,042;**<br>**44.08)** | **Unmet**<br>**(n=38,824 ;**<br>**75.01)** | **Met**<br>**(n=12,896;**<br>**24.99)** | **Unmet**<br>**(n=25,113;**<br>**48.56)** | **Met**<br>**(n=26,607;**<br>**51.44)** |
| **Age** |  |  |  |  |  |  |  |  |  |  |
| 50-54 | 1804(80.04) | 450(19.96) | 1230(54.57) | 1024(45.43) | 574(38.09) | 933(61.91) | 1664(73.82) | 590(26.18) | 881(39.07) | 1374(60.93) |
| 55-59 | 4769(77.48) | 1386(22.52) | 3521(57.21) | 2634(42.79) | 1397(38.24) | 2256(61.76) | 4409(71.63) | 1746(28.37) | 2627(42.68) | 3528(57.32) |
| 60-64 | 6137(70.56) | 2561(29.44) | 4912(56.47) | 3786(43.53) | 1961(39.15) | 3048(60.85) | 6109(70.23) | 2589(29.77) | 3919(45.06) | 4779(54.94) |
| 65-69 | 5759(59.09) | 3987(40.91) | 5400(55.41) | 4345(44.59) | 2374(43.65) | 3065(56.35) | 6794(69.71) | 2952(30.29) | 4517(46.35) | 5229(53.65) |
| 70-74 | 4871(51.17) | 4648(48.83) | 4885(51.32) | 4633(48.68) | 3137(60.06) | 2086(39.94) | 7049(73.97) | 2480(26.03) | 4717(49.56) | 4801(50.44) |
| 75+ | 6786(44.21) | 8562(55.79) | 7432(48.42) | 7916(51.58) | 7100(81.11) | 1654(18.89) | 12809(83.46) | 2539(16.54) | 8452(55.07) | 6896(44.93) |
| **Gender** |  |  |  |  |  |  |  |  |  |  |
| Male | 12688(57.32) | 9447(42.68) | 12357(55.83) | 9778(44.17) | - | - | 16462(74.37) | 5673(25.63) | 11320(51.14) | 10815(48.86) |
| Female | 17438(58.94) | 12147(41.06) | 15023(50.78) | 14562(49.22) | 16543(55.92) | 13042(44.08) | 22362(75.59) | 7223(24.41) | 13793(46.62) | 15792(53.38) |
| **Education** |  |  |  |  |  |  |  |  |  |  |
| Low | 8275(53.59) | 7166(46.41) | 9275(60.07) | 6166(39.93) | 6426(66.50) | 3237(33.50) | 12342(79.93) | 3099(20.07) | 9905(64.15) | 5536(35.85) |
| Medium | 15282(64.91) | 8262(35.09) | 12581(53.44) | 10963(46.56) | 6962(53.51) | 6048(46.49) | 17654(74.98) | 5890(25.02) | 11201(47.57) | 12343(52.43) |
| High | 6569(51.58) | 6166(48.42) | 5524(43.38) | 7211(56.62) | 3155(45.65) | 3757(54.35) | 8828(69.32) | 3907(30.68) | 4007(31.46) | 8728(68.54) |
| **Economic Status** |  |  |  |  |  |  |  |  |  |  |
| Low | 16415(64.93) | 8868(35.07) | 15792(62.46) | 9491(37.54) | 9987(64.50) | 5497(35.50) | 20571(81.36) | 4712(18.64) | 16355(64.69) | 8928(35.31) |
| High | 13711(51.86) | 12726(48.14) | 11588(43.83) | 14849(56.17) | 6556(46.49) | 7545(53.51) | 18253(69.04) | 8184(30.96) | 8758(33.13) | 17679(66.87) |
| **Clinical Diseases** |  |  |  |  |  |  |  |  |  |  |
| Heart | 3291(51.33) | 3120(48.67) | 3323(51.83) | 3088(48.17) | 2040(68.20) | 951(31.80) | 4913(76.63) | 1498(23.37) | 3573(55.73) | 2838(44.27) |
| Hypertension | 13317(53.90) | 11386(46.10) | 12714(51.47) | 11989(48.53) | 8549(60.91) | 5487(39.09) | 18832(76.23) | 5871(23.77) | 12904(52.24) | 11799(47.76) |
| Cholesterol | 7232(49.55) | 7363(50.45) | 7069(48.43) | 7526(51.57) | 4643(55.30) | 3753(44.70) | 10617(72.74) | 3978(27.26) | 7081(48.52) | 7514(51.48) |
| Stroke | 1025(54.26) | 864(45.74) | 935(49.50) | 954(50.50) | 595(67.61) | 285(32.39) | 1418(75.07) | 471(24.93) | 1028(54.42) | 861(45.58) |
| Diabetes | 3954(50.78) | 3833(49.22) | 3492(44.84) | 4295(55.16) | 2616(64.20) | 1459(35.80) | 5894(75.69) | 1893(24.31) | 4420(56.76) | 3367(43.24) |
| Lung | 1482(47.48) | 1639(52.52) | 1470(47.10) | 1651(52.90) | 1004(57.01) | 757(42.99) | 2257(72.32) | 864(27.68) | 1516(48.57) | 1605(51.43) |
| Cancer | 1361(50.24) | 1348(49.76) | 1217(44.92) | 1492(55.08) | 565(37.67) | 935(62.33) | 1751(64.64) | 958(35.36) | 1214(44.81) | 1495(55.19) |
| Ulcer | 900(55.11) | 733(44.89) | 812(49.72) | 821(50.28) | 571(57.39) | 424(42.61) | 1109(67.91) | 524(32.09) | 876(53.64) | 757(46.36) |
| Parkinson's | 192(50.00) | 192(50.00) | 179(46.61) | 205(53.39) | 121(65.41) | 64(34.59) | 295(76.82) | 89(23.18) | 203(52.86) | 181(47.14) |
| Cataracts | 1855(46.04) | 2174(53.96) | 882(21.89) | 3147(78.11) | 1612(64.25) | 897(35.75) | 2985(74.09) | 1044(25.91) | 1773(44.01) | 2256(55.99) |
| Fracture | 1582(56.26) | 1230(43.74) | 1258(44.74) | 1554(55.26) | 1065(59.87) | 714(40.13) | 2109(75.00) | 703(25.00) | 1246(44.31) | 1566(55.69) |
| Dementia | 335(50.00) | 335(50.00) | 346(51.64) | 324(48.36) | 279(75.00) | 93(25.00) | 539(80.45) | 131(19.55) | 416(62.09) | 254(37.91) |
| Psycho | 1678(56.23) | 1306(43.77) | 1367(45.81) | 1617(54.19) | 1145(35.16) | 2112(64.84) | 872(72.49) | 331(27.51) | 1410(47.25) | 1574(52.75) |
| Arthritis | 6593(51.88) | 6115(48.12) | 5909(46.50) | 6799(53.50) | 5154(35.52) | 9356(64.48) | 3352(75.04) | 1115(24.96) | 6018(47.36) | 6690(52.64) |
| Kidney | 536(50.85) | 518(49.15) | 477(45.26) | 577(54.74) | 369(32.77) | 757(67.23) | 297(73.33) | 108(26.67) | 557(52.85) | 497(47.15) |
| **European Countries** |  |  |  |  |  |  |  |  |  |  |
| Austria | 2570(75.48) | 835(24.52) | 1504(44.17) | 1901(55.83) | 1035(51.52) | 974(48.48) | 2719(79.85) | 686(20.15) | 1432(42.06) | 1973(57.94) |
| Germany | 2026(45.35) | 2441(54.65) | 1833(41.03) | 2634(58.97) | 1439(55.80) | 1140(44.20) | 2684(60.10) | 1783(39.90) | 1166(26.10) | 3301(73.90) |
| Netherlands | 184(82.14) | 40(17.86) | 170(75.89) | 54(24.11) | 83(66.94) | 41(33.06) | 184(82.14) | 40(17.86) | 107(47.77) | 117(52.23) |
| Spain | 759(36.37) | 1328(63.63) | 1461(70.00) | 626(30.00) | 731(60.61) | 475(39.39) | 1807(86.61) | 280(13.39) | 1467(70.29) | 620(29.71) |
| Italy | 2097(57.88) | 1526(42.12) | 2215(61.14) | 1408(38.86) | 1384(67.02) | 681(32.98) | 3072(84.76) | 551(15.24) | 2384(65.80) | 1239(35.20) |
| France | 1869(64.25) | 1040(35.75) | 2081(71.54) | 828(28.46) | 886(54.59) | 737(45.41) | 2408(82.78) | 501(17.22) | 2062(70.88) | 847(29.12) |
| Denmark | 1476(62.86) | 872(37.14) | 1273(54.22) | 1075(45.78) | 925(63.18) | 539(36.82) | 1905(81.13) | 443(18.87) | 1365(58.13) | 983(41.87) |
| Greece | 1680(53.98) | 1432(46.02) | 2255(72.46) | 857(27.54) | 1277(71.98) | 497(28.02) | 2734(87.85) | 378(12.15) | 2385(76.64) | 727(23.36) |
| Switzerland | 857(46.48) | 987(53.52) | 615(33.35) | 1229(66.65) | 513(51.87) | 476(48.13) | 1261(68.38) | 583(31.62) | 313(16.97) | 1531(83.03) |
| Belgium | 2266(50.39) | 2231(49.61) | 1933(42.98) | 2564(57.02) | 1134(44.59) | 1409(55.41) | 2874(63.91) | 1623(36.09) | 1457(32.40) | 3040(67.60) |
| Israel | 228(30.65) | 516(69.35) | 289(38.84) | 455(61.16) | 206(50.99) | 198(49.01) | 639(85.89) | 105(14.11) | 142(19.09) | 602(80.91) |
| Czech Republic | 2021(60.89) | 1298(39.11) | 1247(37.57) | 2072(62.43) | 926(50.11) | 924(49.89) | 2373(71.50) | 946(28.50) | 717(21.60) | 2602(78.40) |
| Luxembourg | 612(75.56) | 198(24.44) | 497(61.36) | 313(38.64) | 256(53.11) | 226(46.89) | 492(60.74) | 318(39.26) | 450(55.56) | 360(44.44) |
| Hungary | 1612(88.18) | 216(11.82) | 1202(65.75) | 626(34.25) | 755(71.27) | 304(28.73) | 1659(90.75) | 169(9.25) | 1364(89.39) | 464(10.61) |
| Estonia | 1945(43.36) | 2541(56.64) | 1948(43.42) | 2538(56.58) | 1198(48.38) | 1279(51.62) | 3309(73.70) | 1177(26.30) | 2001(44.61) | 2485(55.39) |
| Croatia | 2807(59.89) | 1880(40.11) | 2643(56.39) | 2044(43.61) | 1679(61.75) | 1039(38.25) | 3611(77.04) | 1076(22.96) | 2735(58.35) | 1952(41.65) |
| Lithuania | 777(54.95) | 637(45.05) | 720(50.82) | 694(49.18) | 395(50.32) | 390(49.68) | 997(70.51) | 417(29.49) | 518(36.63) | 896(63.37) |
| Bulgaria | 442(53.12) | 389(46.88) | 334(40.19) | 497(59.81) | 245(51.26) | 233(48.74) | 601(72.32) | 230(27.68) | 259(31.17) | 572(68.83) |
| Cyprus | 558(75.82) | 178(24.18) | 267(36.28) | 469(63.72) | 173(37.94) | 283(62.06) | 466(63.32) | 270(36.68) | 228(30.98) | 508(69.02) |
| Finland | 1295(73.66) | 463(26.34) | 1243(70.71) | 515(29.29) | 493(49.75) | 495(40.25) | 1344(76.45) | 414(23.55) | 1204(68.49) | 554(31.51) |
| Latvia | 1352(78.79) | 364(21.21) | 1042(60.72) | 674(39.28) | 524(51.07) | 502(48.93) | 1031(60.08) | 685(39.92) | 929(54.14) | 787(45.86) |
| Malta | 693(79.20) | 182(20.80) | 608(69.49) | 267(30.51) | 286(58.85) | 200(41.15) | 654(74.74) | 221(25.26) | 428(48.91) | 447(51.09) |

### Preventive Care-specific Predictive Model Performance

LightGBM outperformed all other machine learning algorithms across all five preventive care outcomes, assessed by ROC curves and AUC value (Figure 1). By various key preventive care outcomes, with AUC performance of 0.81 for dental care in LightGBM and GradientBoosting model, 0.78 for colorectal cancer screening, 0.74 for mammogram and eye examination, and 0.73 for vaccination. Compared with logistic regression models, LightGBM yielded absolute AUC improvements of 0.05–0.06 (a relative gain of 7–8%) Receiver operating characteristic (ROC) curves confirms that LightGBM model performs consistently high accuracy across five preventive care services.

**Figure 1.**
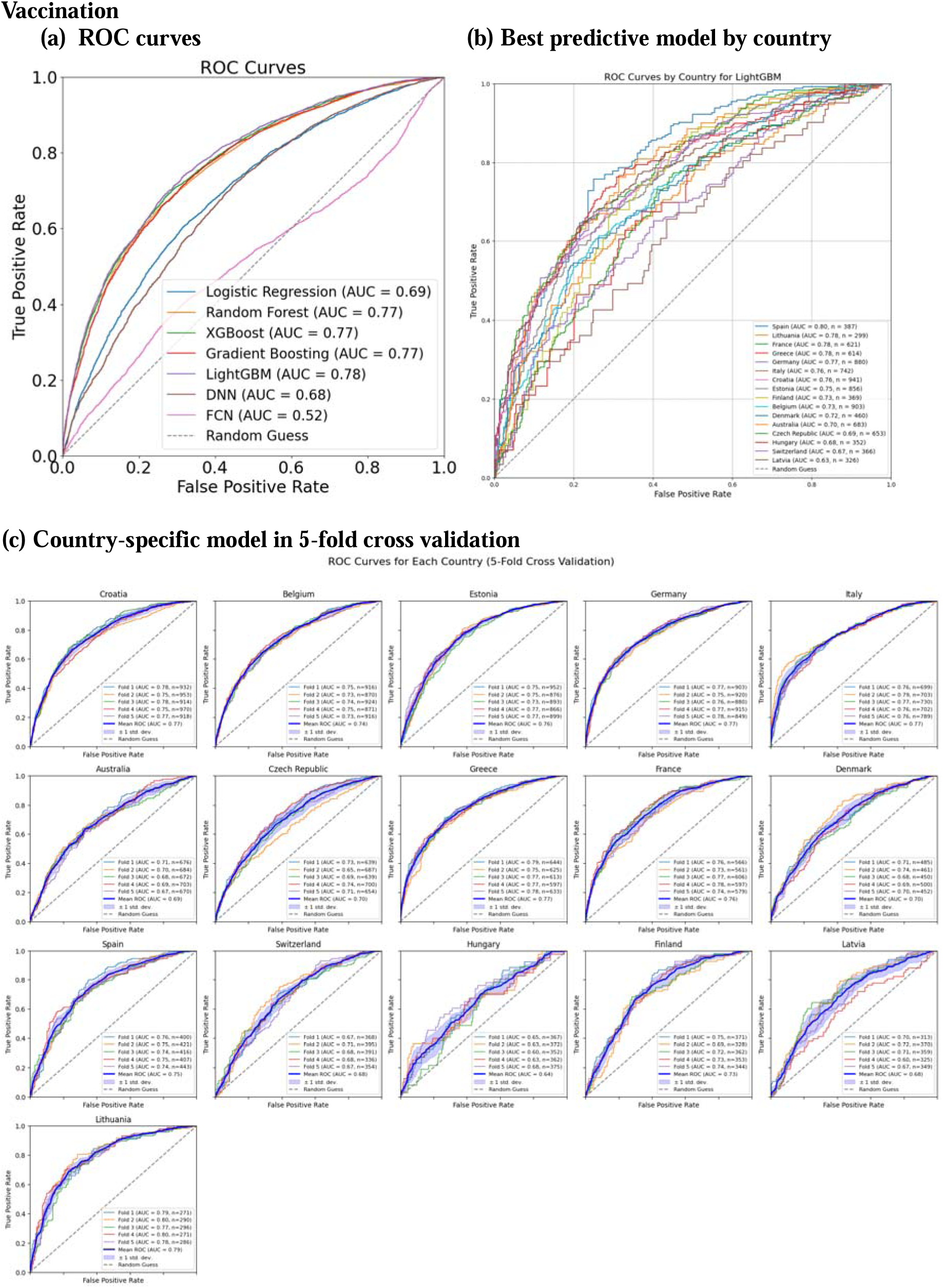
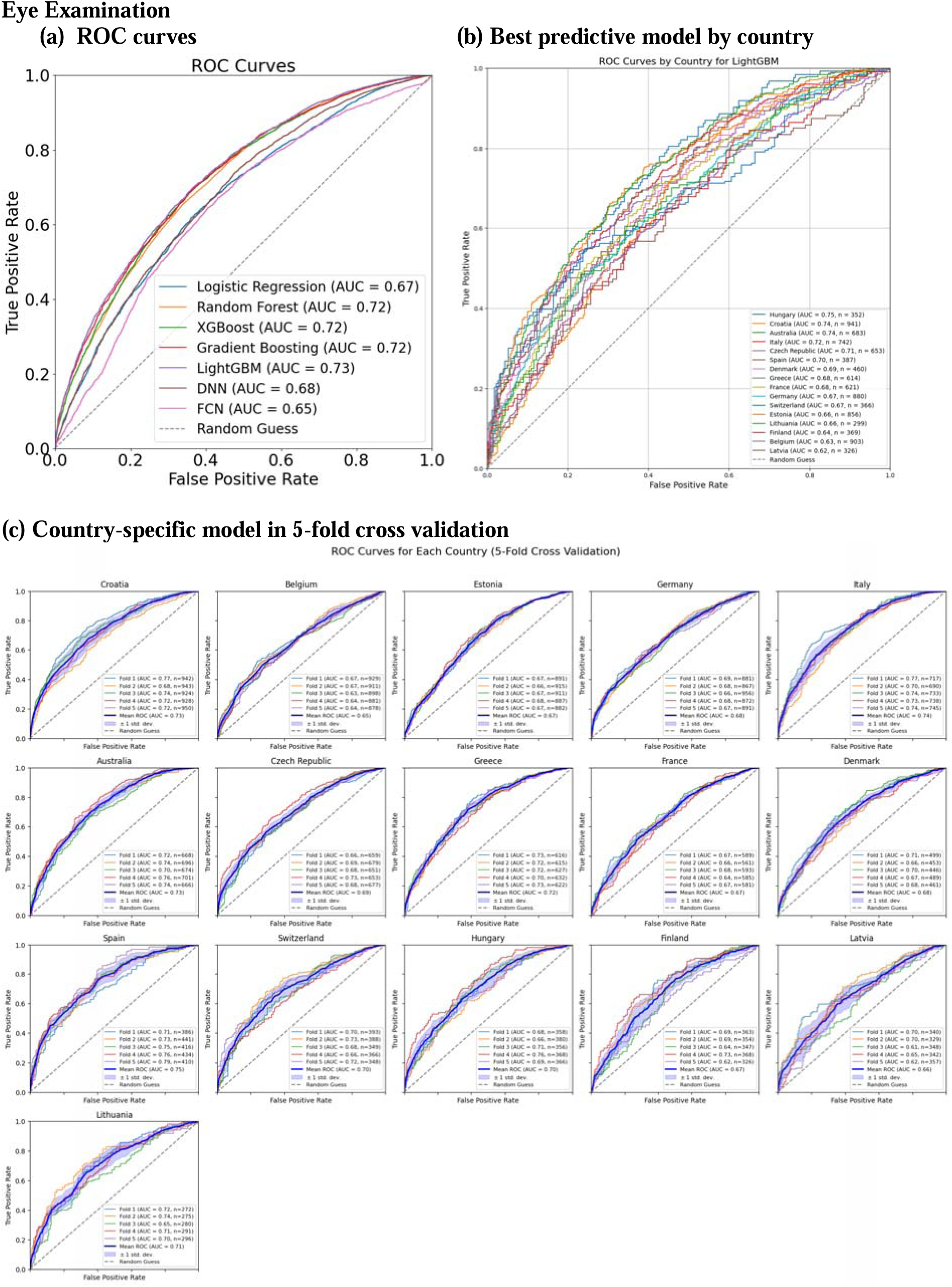
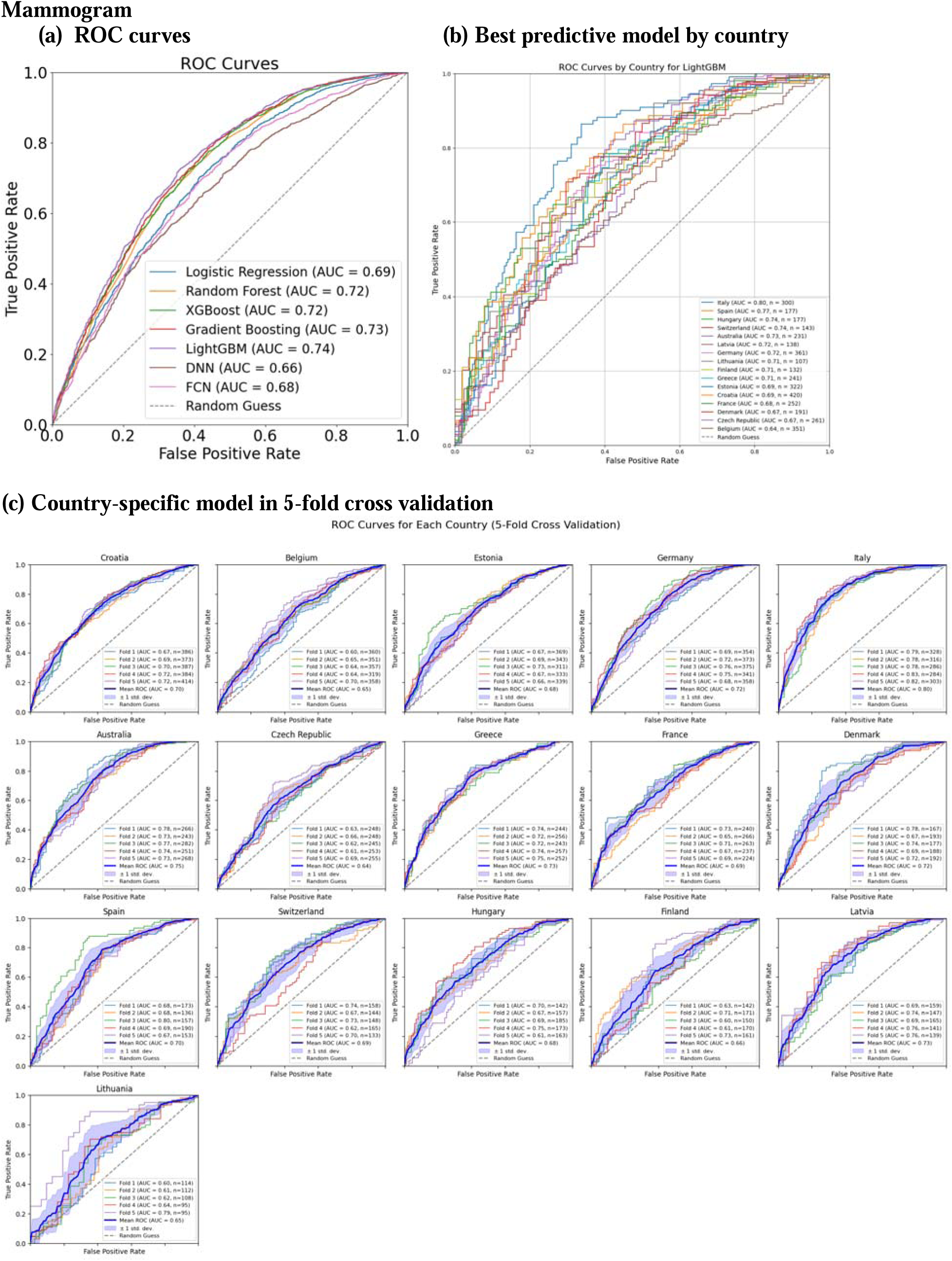
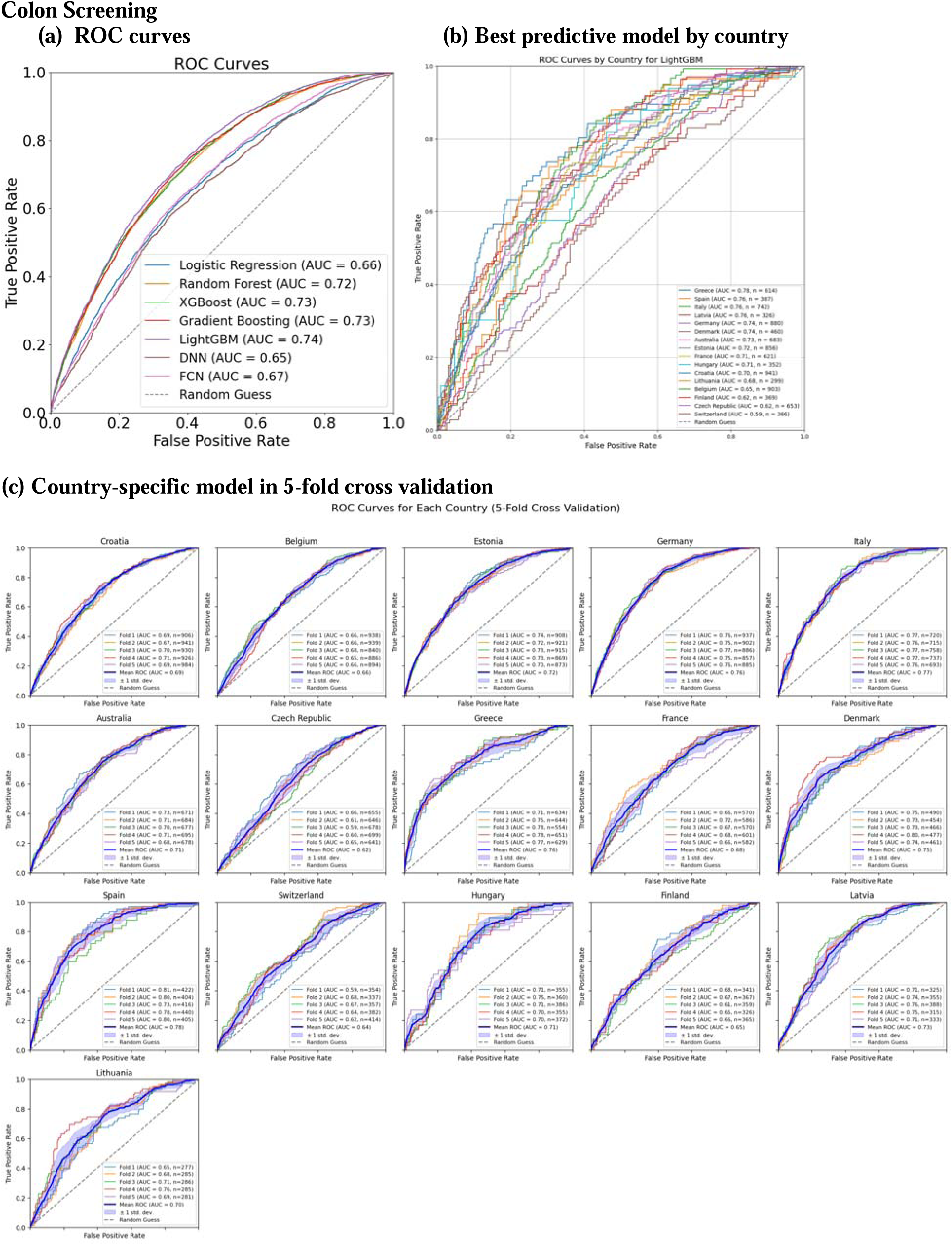
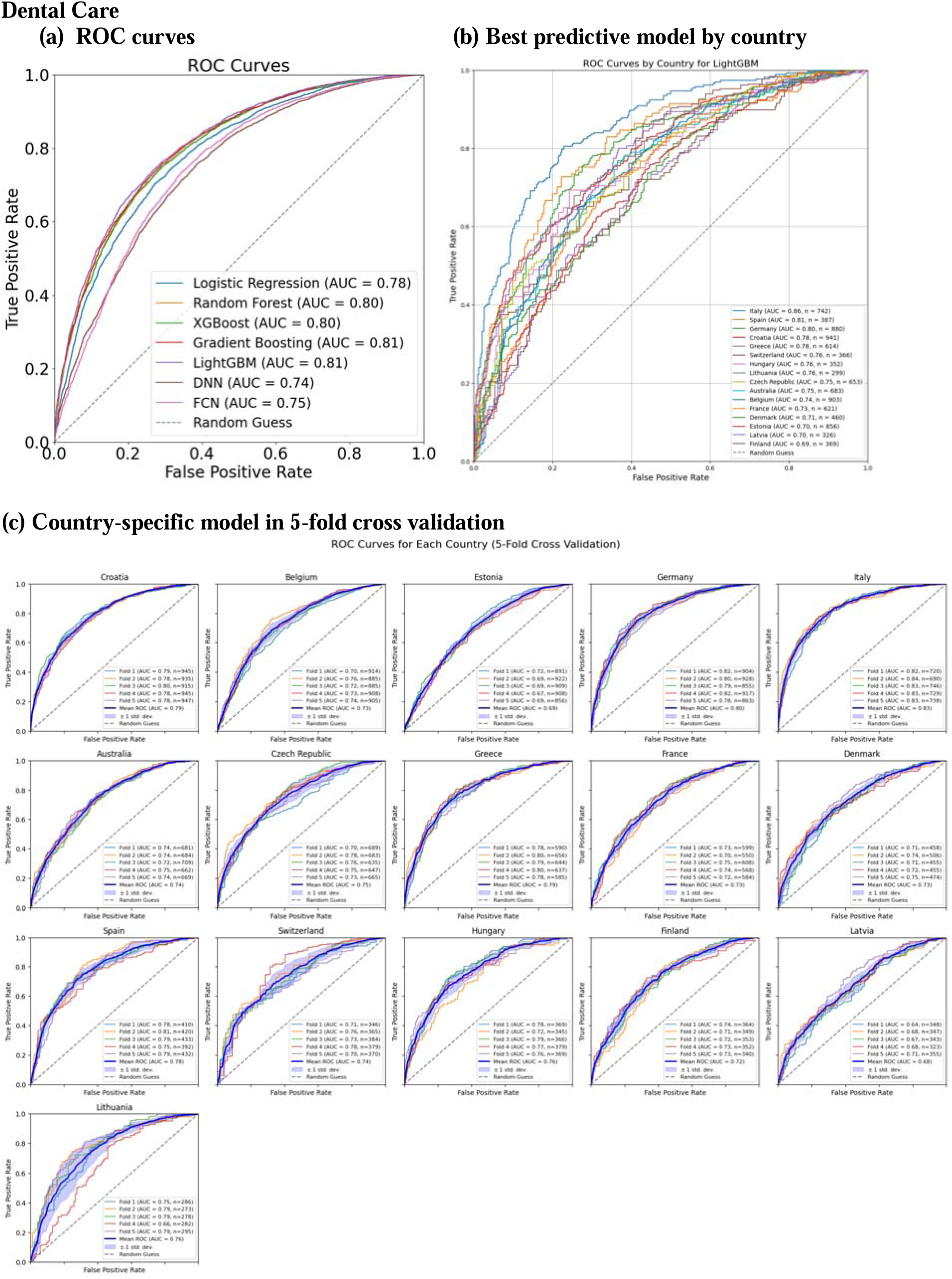
ROC Curves Vaccination.

### Country-specific Predictive Model Performance

In country specific machine learning model (Figure 1), LightGBM model’s discriminatory power varied across preventive care services. For vaccination, Spain achieved the highest AUC (0.80), while Hungary led in predicting unmet needs for eye examinations (AUC = 0.75). Italy demonstrated the strongest performance for mammograms (AUC = 0.80) and dental care (AUC = 0.86), whereas Greece topped colon cancer screening (AUC = 0.78). In contrast, Switzerland’s models performed comparatively poorly, yielding AUC below 0.70 for several outcomes, highlighting regional variations.

### Predictive Model Performance across subgroups

The heat maps (Figure 2) reveals the differences of predictive model performance across European countries and socioeconomic subgroups for the five key preventive care services. For vaccination, LightGBM attained highest disparity in Italy (AUC = 0.78), whereas the FCN performs lowest AUC of 0.57 in Switzerland. In eye examination, LightGBM achieves highest AUC of 0.75 in Spain and the DNN model underperformed most in Belgium (AUC = 0.63).

**Figure 2.**
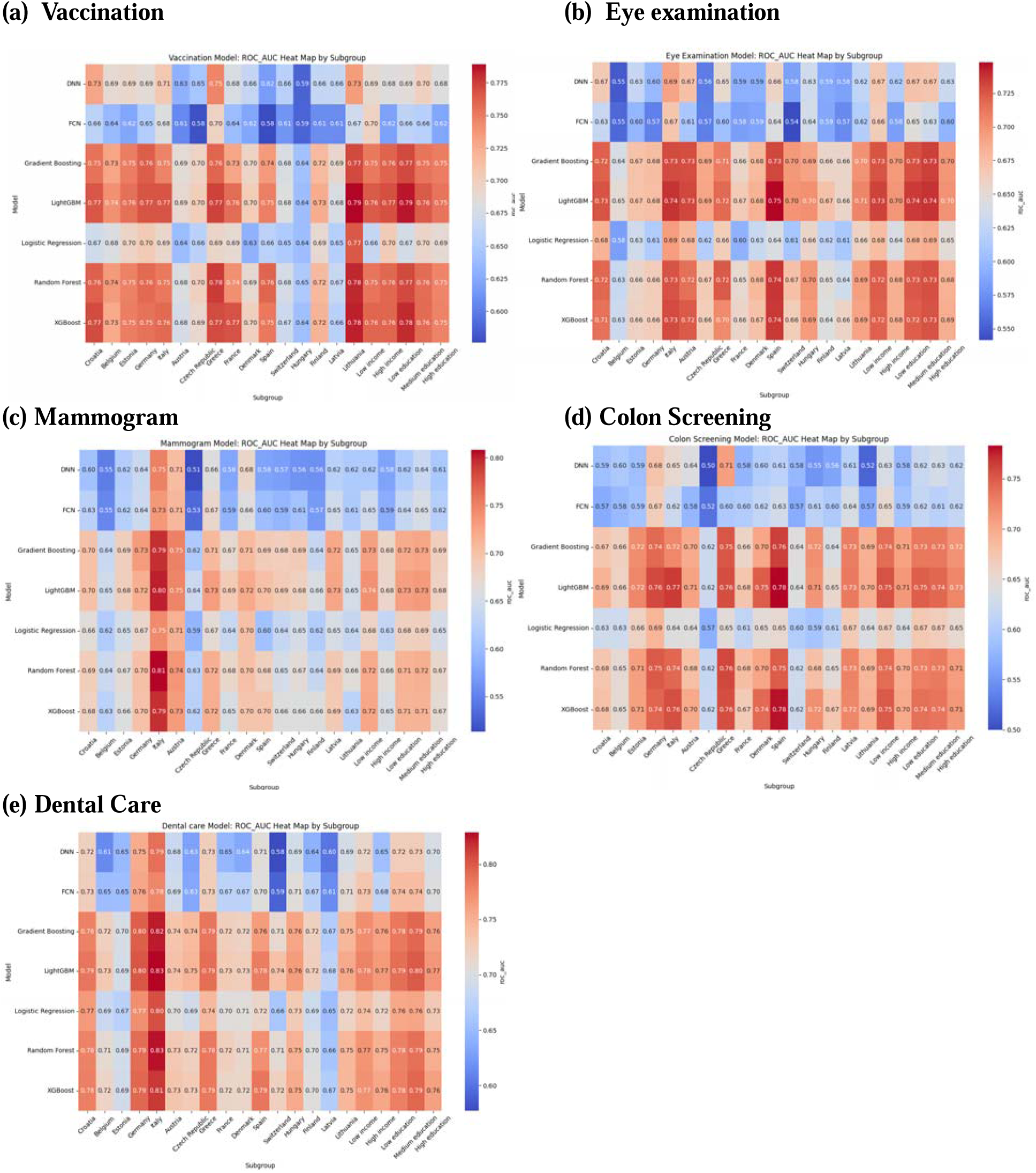
Heat map: Model performance AUC by Socioeconomic and European countries subgroups.

Mammography was best predicted by LightGBM in Italy (AUC = 0.81), while the DNN fell to 0.50 in the Czech Republic. For colon screening, Gradient Boosting performed best in Greece (AUC = 0.77) and worse in the Czech Republic by FCN model (AUC = 0.48). For dental care, LightGBM excel in Italy (AUC = 0.83) with the FCN weakest in the Czech Republic (AUC = 0.59). Across socioeconomic subgroups, LightGBM consistently outperformed deep learning models, especially among medium education participants (AUC = 0.70-0.76), underscoring the need for locally tailored predictive tools in preventive medicine.

### Fairness Evaluation

We evaluated algorithmic fairness of the LightGBM model across the five preventive care outcomes using two established fairness metrics: demographic parity and equalized odds, the latter assessed through both true positive rate (TPR) and false positive rate (FPR). These metrics were examined across 16 European countries and within key socioeconomic subgroups defined by income and education levels. Detailed results are presented in Table 2 and visualized in Figure 2.

**Table 2.**
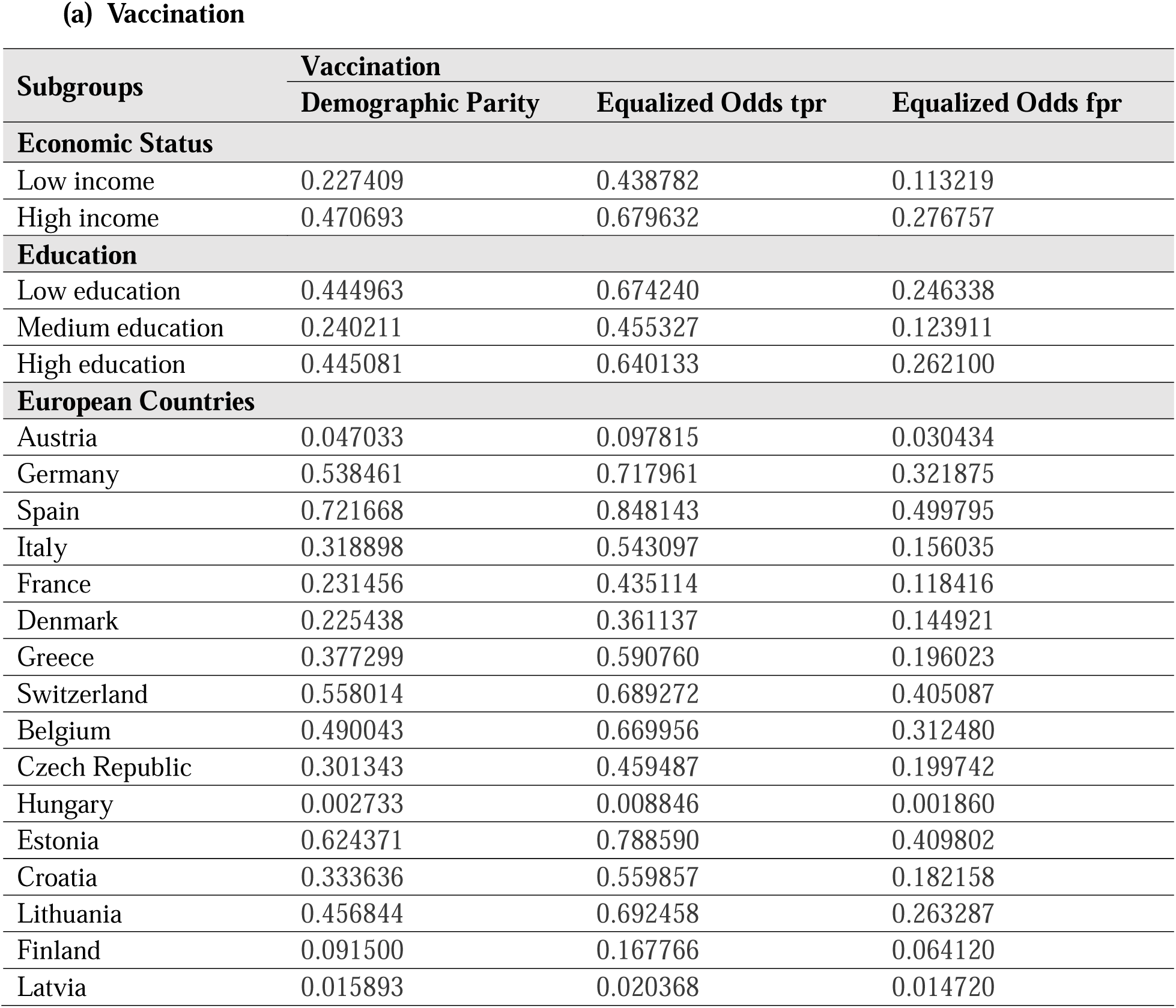
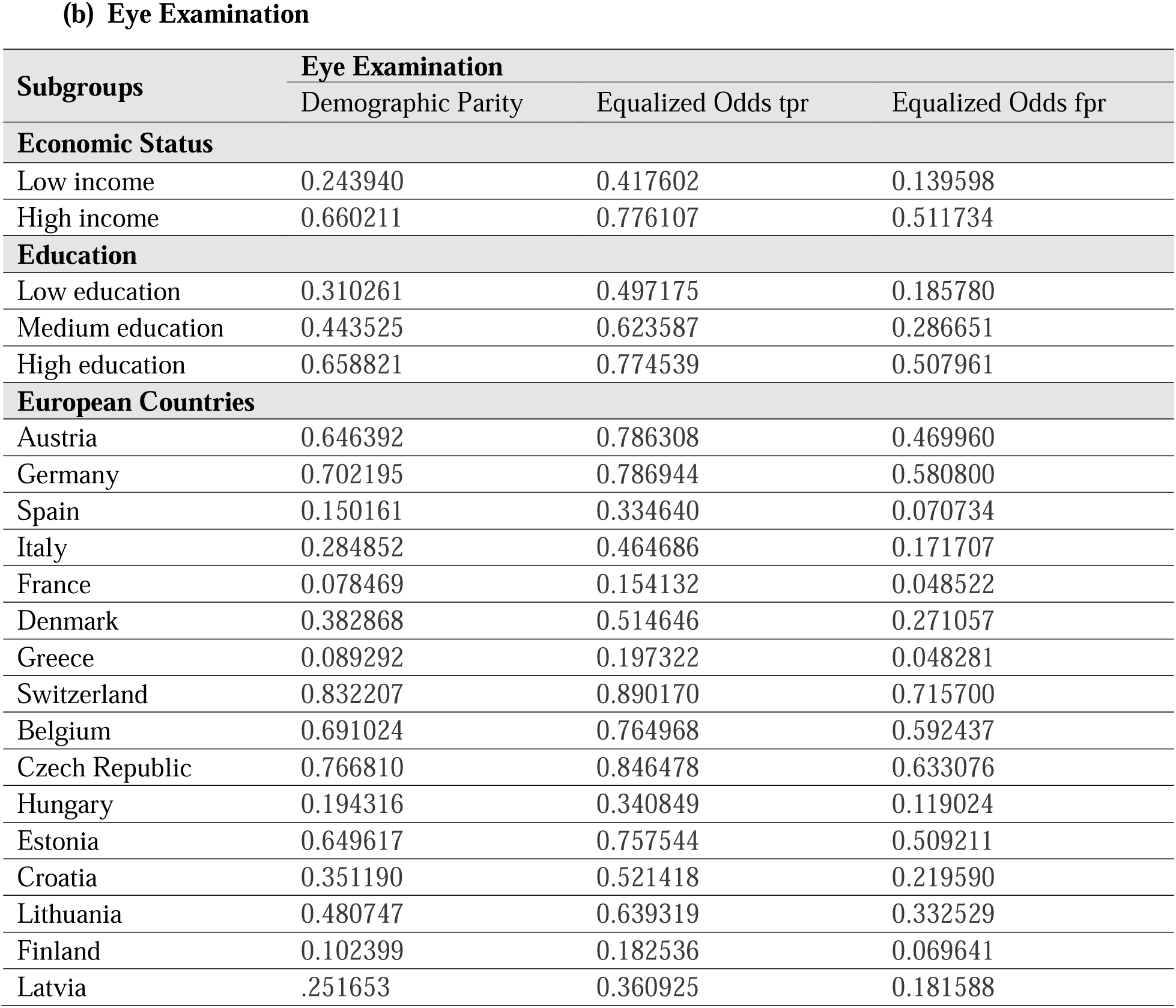
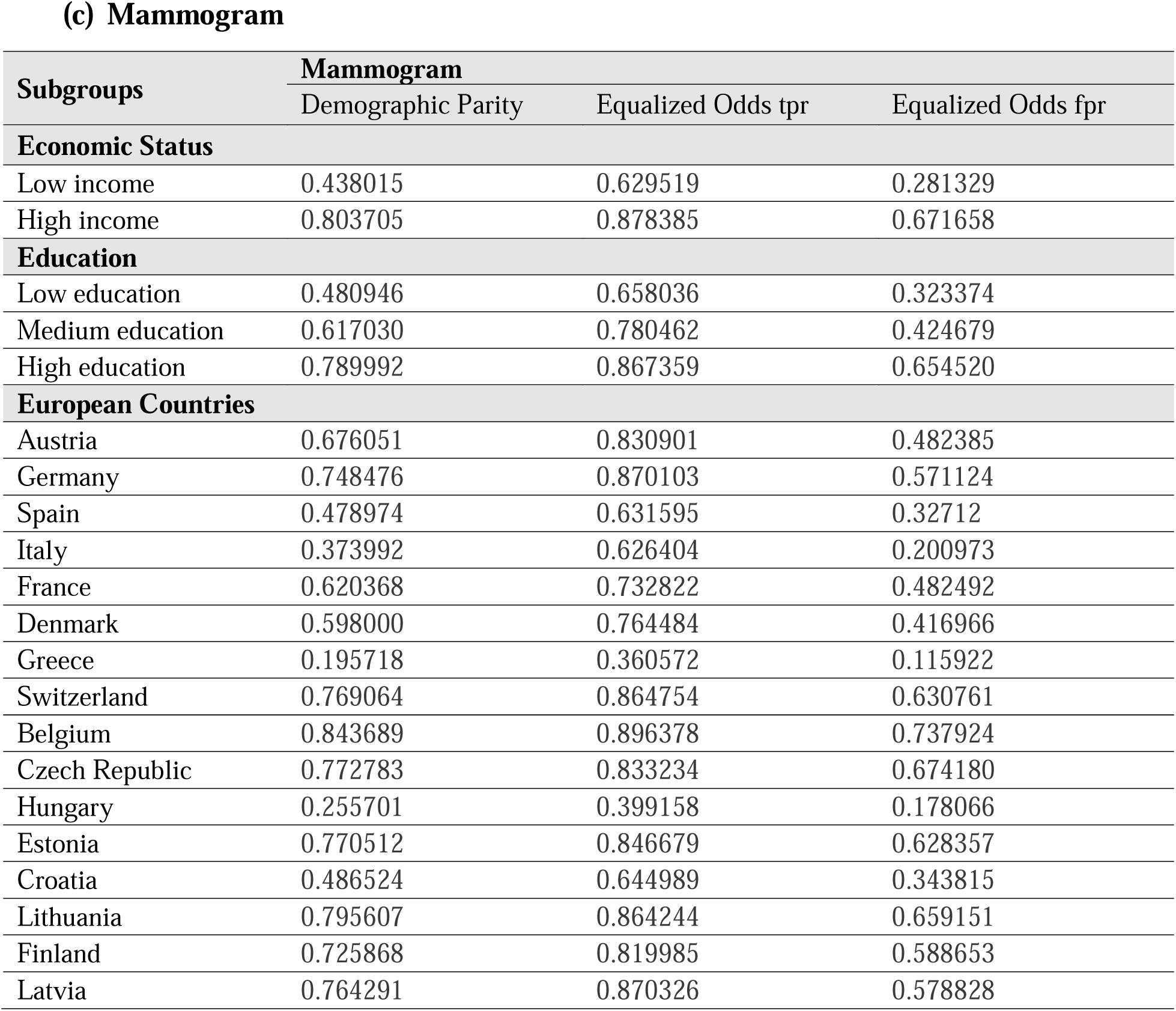
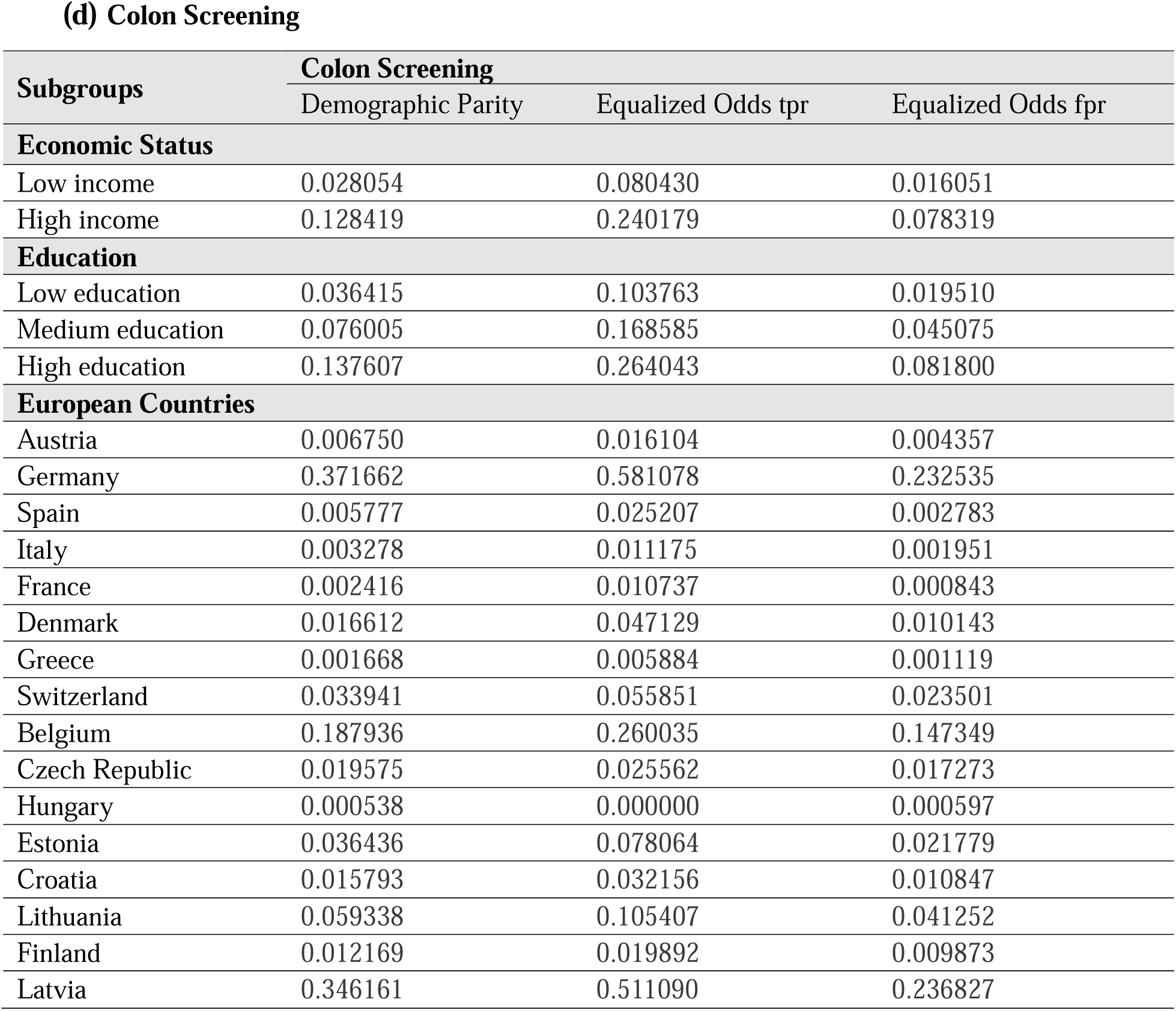
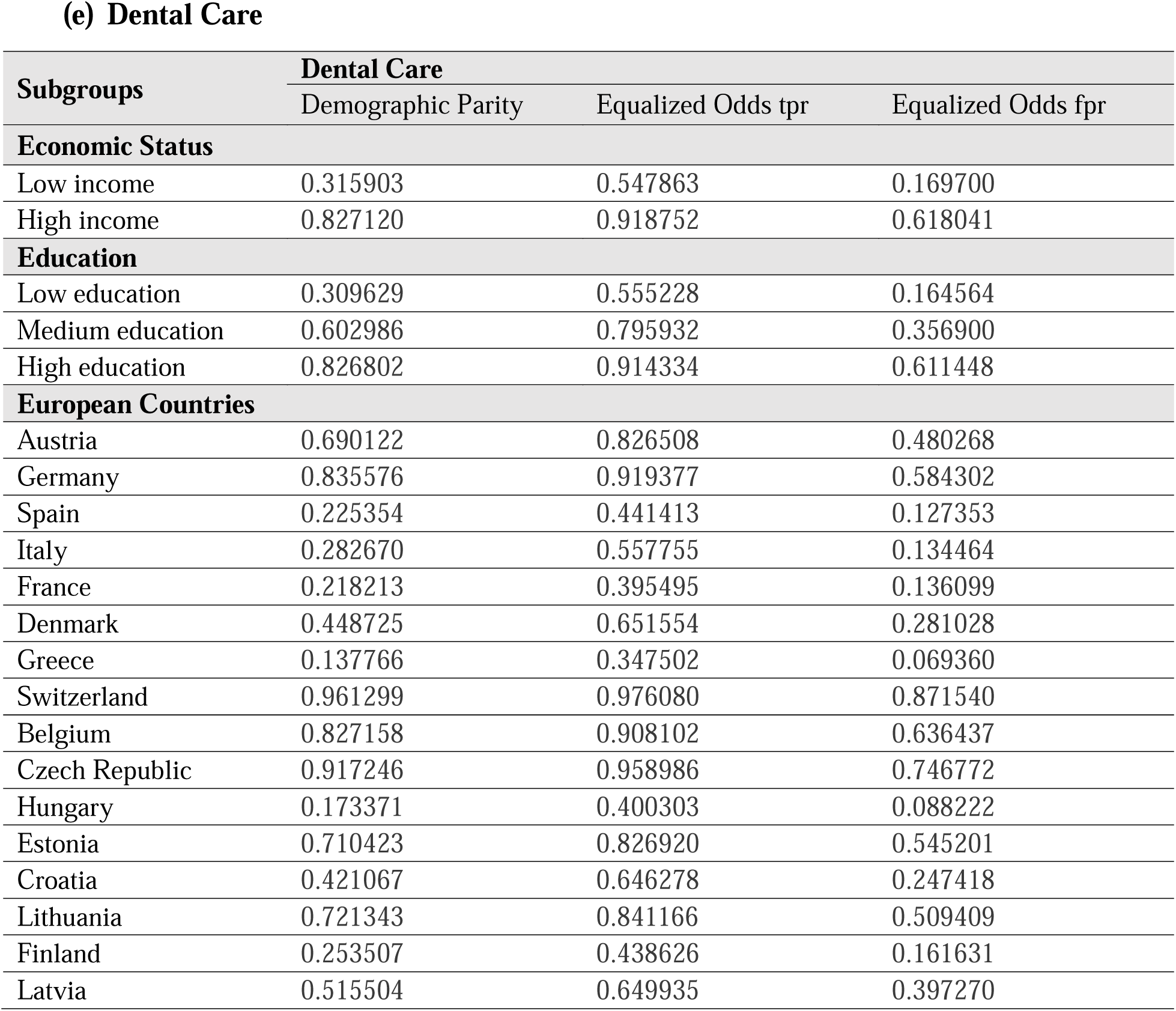
Fairness Metrics of Preventive Care across Socioeconomic and European Countries Subgroups for Best Model.

Substantial variability in fairness metrics was observed across both geographic and socioeconomic dimensions. For dental care, demographic parity values ranged widely across countries, from 0.14 in Greece to 0.96 in Switzerland, while equalized odds–FPR ranged from 0.07 to 0.87. Similarly, in the case of eye examination, Switzerland exhibited the highest demographic parity at 0.83 and the highest FPR at 0.72, whereas France demonstrated much lower values, with demographic parity of 0.08 and FPR of 0.05. For vaccination, Hungary showed the lowest demographic parity (0.003) and TPR (0.009), while Estonia reached the highest levels (0.62 for demographic parity and 0.79 for TPR), indicating considerable disparity in how the model performed across national contexts.

Mammogram predictions also revealed pronounced disparities. Belgium and Estonia had the highest demographic parity values, both exceeding 0.77, while Greece reported the lowest at 0.20. In colon screening, fairness disparities were comparatively smaller, with demographic parity ranging from 0.0005 in Hungary to 0.35 in Latvia. Across all five outcomes, country-specific equalized odds FPR frequently exceeded 0.60 in several settings, particularly in Switzerland, Germany, and Belgium, suggesting that some models had high false positive error rates in specific countries despite overall good discrimination performance.

Fairness across socioeconomic subgroups also demonstrated notable differences. For eye examinations, the high-income group showed a demographic parity of 0.66 and FPR of 0.51, in contrast to the low-income group, which had respective values of 0.24 and 0.14. Among education subgroups, high-education individuals exhibited higher fairness disparities across most outcomes. In the mammogram outcome, the demographic parity for the high-education group reached 0.79, whereas it was 0.48 for the low-education group. For dental care, high-income individuals had a demographic parity of 0.83 and FPR of 0.62, significantly higher than the corresponding values of 0.32 and 0.17 for the low-income group.

Across all outcomes, the equalized odds–TPR and FPR metrics generally exhibited greater variability than demographic parity, highlighting inconsistencies in model sensitivity and error distribution. These discrepancies were particularly evident in countries with high predictive performance, such as Italy and Germany, where fairness gaps remained substantial. The full range of AUC and fairness metrics by country and subgroup is displayed in Figure 2, which provides a comparative heatmap overview of how model fairness varies by geography and social determinants.

### SHAP and Features Importance

SHAP analysis of the optimal LightGBM predictive model (Figure 3) identified key predictors of unmet preventive care needs, with social engagement frequency, socioeconomic status, age, quality of life (CASP-12), supplementary insurance coverage, and outpatient emerging as principal contributors. For vaccination, the most influential predictors were better quality of life (SHAP value approximately 0.23) and frequency of social engagement (0.19), followed by age (0.15). In the context of eye examination, social engagement frequency achieves the greatest impact (0.31), along with outpatient visits (0.11) and quality of life (0.09). Predictors of mammogram includes social frequency (0.15), supplementary insurance (0.13), and economic status (0.11). For colon screening, age (0.33) and social engagement (0.29) were most influential, followed by supplementary insurance (0.12). Dental care was primarily predicted by prior eye examination (0.33), mammography history (0.24), and economic status (0.16). These findings emphasize the consistent relevance of demographic and socioeconomic factors while illustrating the procedure-specific contributions of social support networks and prior preventive care behaviors.

**Figure 3.**
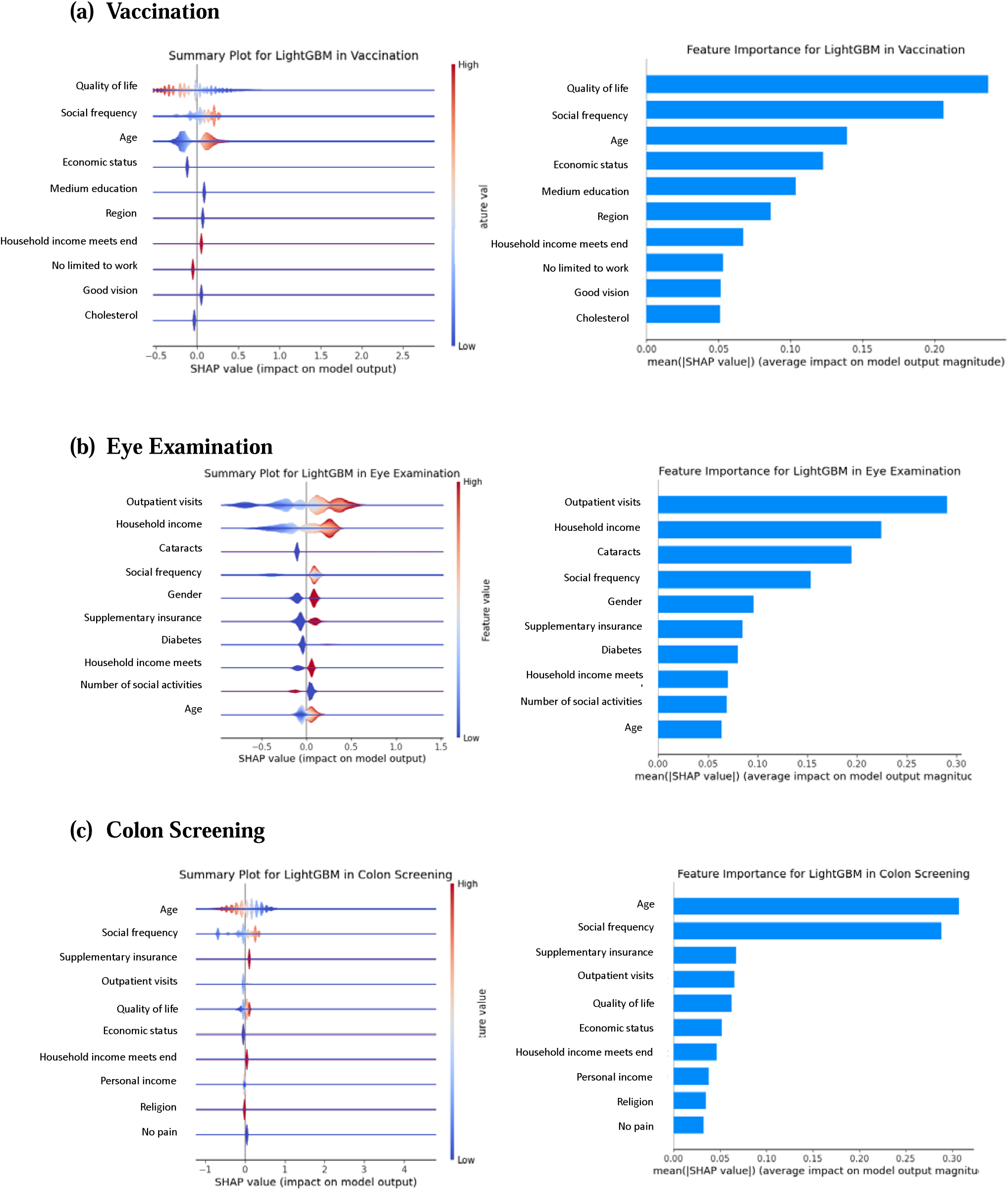
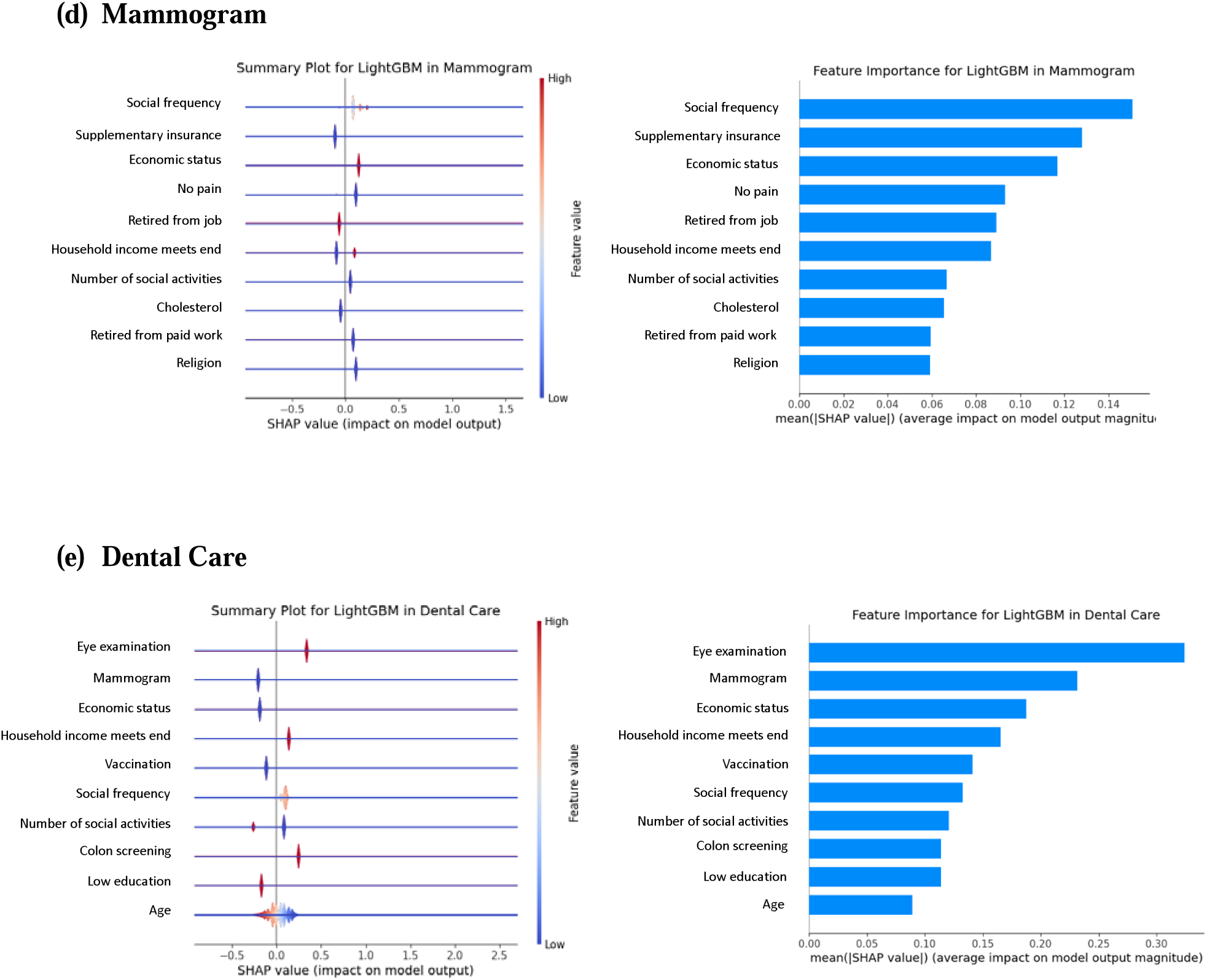
SHAP and Feature Importance Analysis.

## DISCUSSION

In this cross-national study using data from 51,720 older adults across 16 European countries, we developed and evaluated machine learning models to predict five categories of unmet preventive care needs. LightGBM consistently achieved the highest overall predictive performance across all outcomes, with AUCs ranging from 0.73 for eye examination to 0.81 for dental care.

However, a central and novel finding of this study is that predictive performance alone does not guarantee fairness of ML model.

Substantial disparities in fairness were identified both across countries and within socioeconomic subgroups. Even when discrimination metrics such as AUC were high, fairness evaluations revealed large differences in model behavior. For example, demographic parity for dental care predictions ranged from 0.137766 in Greece to 0.961299 in Switzerland, while equalized odds false positive rates exceeded 0.80 in several countries. Such discrepancies suggest that the same model may systematically over- or under-identify unmet needs depending on geographic context.

Socioeconomic subgroups exhibited similar divergence. In the eye examination model, the high-income group had a demographic parity of 0. 0.660211 and a false positive rate of 0.511734, compared to 0.243940 and 0.139598 in the low-income group, respectively. Fairness disparities were consistently more pronounced in high-income and high-education subgroups, despite the model being trained on pooled European data.

These findings underscore that algorithmic fairness is not inherently achieved by high-performing models. Even well-calibrated algorithms such as LightGBM can yield unequal performance across social and geographic subgroups if fairness is not explicitly addressed. Therefore, evaluating subgroup-specific fairness metrics is critical before deploying machine learning tools for population-level preventive health planning, particularly in multinational or heterogeneous settings.

Unmet preventive care needs remain a substantial public health concern in Europe, with persistent gaps in vaccination, cancer screening, and dental care disproportionately affecting older adults and socially disadvantaged populations.^23^ ^24^ Our findings reinforce previous evidence that underutilization of preventive services including vaccination, colon screening, and dental care persists even in high-resource settings, reflecting complex interactions between individual, social, and systemic determinants.^1^ ^25^ These disparities not only contribute to avoidable morbidity and mortality, but also signal persistent challenges in achieving equitable population health outcomes.^26^

Machine learning (ML) models, when rigorously evaluated and appropriately applied, provide new opportunities for public health surveillance and resource targeting in preventive care. By harnessing multidimensional data on demographic, socioeconomic, and clinical factors, ML enables identification of at-risk subgroups who are likely to experience gaps in preventive service uptake.^27^ However, our results indicate that the predictive performance of ML models can differ notably across countries and social subgroups, and high discrimination does not equate to fair or equitable predictions.^9^ Fairness evaluations reveal that subgroups especially those with higher education or income may benefit unequally from model-based predictions, while more vulnerable groups continue to face under-identification or misclassification.^12^

This study highlights the importance of embedding fairness considerations within the development and assessment of ML models intended for public health applications. Addressing disparities in unmet preventive care is central to the goal of improving population health and reducing health inequalities.^28^ Model evaluation should extend beyond accuracy metrics to include rigorous assessments of subgroup fairness, particularly when models are applied in diverse, multinational contexts.^29^

### Policy and Research Implications

Our findings underscore the need for fairness aware, country specific ML models in preventive cares. We recommend the following action to ensuring ML model applied in healthcare and preventive cares. (1) Funding pilot programs that incorporate demographic parity and equalized odds constraints into national screening algorithms; (2) Mandating independent fairness audits reporting subgroup metrics alongside overall performance prior to public procurement of predictive tools; (3) Creating interdepartmental task forces (digital, health, statistics) to harmonize data standards and develop equitable ML guidelines; and (4) Upgrading data systems to capture key social determinants (e.g., education, rural and urban residence) for more granular subgroup analyses.

Policymakers must prioritize investments in robust data infrastructure and disaggregated data collection to support subgroup-specific analyses, reduce biases, and enhance resource allocation. Embedding fairness-focused ML models into public health policies can effectively address health inequities and set a global standard for ethical AI in healthcare. Moreover, our pooled data design may obscure intra country heterogeneity, warranting future studies with finer geospatial and contextual data.

## CONCLUSION

This study highlights the importance of fairness-aware ML models in addressing preventive care disparities across European countries. LightGBM model showed the strongest predictive accuracy, but geographic variations emphasize the need for country-specific subgroup modeling. To leverage the effectiveness and fairness of model, ML models tailors to local contexts should guide policy decisions to reduce inequities, enhance resource allocation, and support universal health coverage.

## Data Availability

All data produced are available online at: https://share-eric.eu/data/

